# HIV pre-exposure prophylaxis among people initiating buprenorphine for opioid use disorder: a retrospective cohort study

**DOI:** 10.1101/2025.11.26.25340877

**Authors:** Alyssa S. Tilhou, Jiayi Wang, Sruthi Treasa George, Laura White, Sabrina A. Assoumou

## Abstract

**Introduction:** Buprenorphine (BUP) is an effective treatment for opioid use disorder (OUD) that may create opportunities to expand access to preventive services for patients with OUD when implemented in the primary care setting. HIV prevention, particularly pre-exposure prophylaxis (PrEP), represents a vital service for persons with OUD due to HIV risk associated with injection drug use and condomless sex. Evaluating the existing integration of BUP treatment and HIV prevention is an important step to further optimize comprehensive care for patients with OUD.

**Methods:** This retrospective cohort study used pharmacy claims from the Merative™ MarketScan^®^ Research Databases, 2014 – 2022, to examine use of PrEP among new BUP treatment episodes over the first 63 days of treatment. We excluded BUP episodes from individuals with probable HIV (based on diagnosis or medications indicated for HIV in the past 365 day). Clinical indications for PrEP were identified based on past 365-day diagnosis of sexually transmitted infections (STIs) or serious injection-related infections (SIRIs).

**Results:** Of 123,740 BUP episodes representing 76,377 individuals, 104 (0.1%) involved PrEP during BUP treatment. Most of these episodes (71%) involved PrEP in the 30 days prior to BUP initiation. Only 6% of the sample exhibited diagnoses indicating risk for HIV. Of these, only 0.2% involved PrEP during the BUP treatment episode. BUP episodes involving PrEP tended to be disproportionately male (89% compared with 63%) and from the Northeast (27% vs. 18%) or West (30% vs. 16%).

**Conclusions:** Findings from this retrospective cohort study identify a major gap in PrEP use for individuals with OUD and documented HIV risk. Aligning HIV prevention and BUP treatment is an important opportunity to enhance health for patients with OUD.

## Introduction

Opioid use disorder (OUD) is a major public health concern affecting over 9 million people in the United States.^1^ Treatment with evidence-based medications for OUD (MOUD) is an essential intervention to reduce risk of opioid overdose. Yet, the health consequences of OUD extend beyond opioid overdose. OUD increases risk for psychiatric comorbidities, infectious diseases, and preventable acute and chronic conditions.^3–5^ MOUD treatment represents a path to improve health in these parallel domains. By reducing nonprescribed opioid use and opioid withdrawal, MOUD treatment enables patients to pursue other health goals.

In particular, OUD treatment in the primary care setting positions patients to access comprehensive health care services. Most primary care-based MOUD treatment involves the medication, buprenorphine (BUP).^6–8^ As such, ambulatory initiation of BUP represents a pivotal opportunity to deliver high-impact interventions to optimize health.

Prevention of human immunodeficiency virus (HIV) is one such priority intervention in the primary care setting.^9,10^ Many patients with OUD experience elevated risk of HIV in association with behaviors such as injection drug use or condomless sex.^11,12^ Thus, promoting HIV prevention when initiating BUP is an important strategy to optimize health. Pre-exposure prophylaxis for HIV (PrEP) offers a particularly effective^13^ and underutilized strategy that integrates well with BUP treatment in primary care settings.^10,14^ In optimal circumstances, patients presenting for BUP treatment would be offered laboratory testing for HIV, assessment of HIV risk factors and initiation of PrEP in line with clinical guidelines.^15^ Thus, not only is PrEP an important health promotion strategy, but it is also a measure of health system performance on behalf of patients with OUD.^10^

Population studies suggest notable underutilization of PrEP among eligible patients, including those with OUD and injection drug use.^16,17^ Some studies suggest that fewer than 1% of individuals eligible for PrEP in association with injection drug use have received PrEP.^17,18^ Whether this underutilization is overcome by engaging in OUD services, such as BUP treatment, is unknown. To understand the role that BUP initiation may play in facilitating PrEP utilization, the current study describes PrEP utilization in the first two months of BUP treatment.

## Methods

### Data

This retrospective cohort study used data from the Merative™ MarketScan^®^ Research Databases, 2014 – 2022. (This time period reflects the FDA approval windows for three PrEP formulations: tenofovir disoproxil fumarate plus emtricitabine in July 2012; tenofovir alafenamide plus emtricitabine in October 2019; and injectable cabotegravir in January 2021.) We used administrative data to assess enrollment continuity and pharmacy claims to examine medication utilization over 63 days (nine weeks) of BUP treatment. We selected a 63-day treatment window in which to observe PrEP to accommodate: 1) patients’ competing needs, which might support initiating PrEP after BUP initiation is complete, and 2) the time required to complete laboratory tests and schedule follow up for PrEP prescribing. Data preparation, including linkage, de-identification, and cleaning were overseen by Merative™ prior to data access and utilization. As such, ethics committee approval was not sought as this study did not constitute human subjects research.

### Cohort

The data were structured as 63-day person-episodes from individuals 16-89 years on Day 1 of BUP treatment. Contributing individuals were identified based on pharmacy claims for a BUP formulation indicated for OUD between 07/01/2015 and 10/30/2022. We then applied the following restrictions: 1) continuous enrollment for 30 days prior to Day 1 of BUP without a claim for BUP in this pre-period, and 2) 63 days of continuous enrollment starting on Day 1 of BUP. We excluded episodes from individuals with HIV based on observing either: 1) an outpatient claim for a diagnosis of HIV (based on the International Classification of Diseases, Ninth Revision (ICD-9) or Tenth Revision (ICD-10)) or 2) an antiretroviral medication indicated for HIV in the 365 days prior to Day 1 of BUP or on Days 1-63 of BUP treatment (see Appendix). We also excluded all subsequent episodes from these individuals. This antiretroviral medication exclusion also served to prevent misclassifying PrEP initiation as hepatitis B (HBV) treatment or post-exposure prophylaxis for HIV. Of note, we did allow individuals with past 365-day diagnoses of HBV to remain eligible for PrEP given risk of acquiring HIV.

### Measures

The primary outcome of interest was receipt of PrEP during the 63-day BUP treatment period based on pharmacy claims. We used days supplied to extrapolate days covered by each PrEP prescription. This approach allowed PrEP prescriptions to count towards utilization during BUP treatment if covered days crossed BUP Day 1. No records for injectable PrEP were observed during the sample of BUP treatment episodes or in the 180 days prior.

Recent PrEP use was defined as observing PrEP receipt in the 30 days prior to BUP initiation (following the same procedures for extrapolating days covered based on pharmacy claims). HIV risk was assessed based on ICD-9 and ICD-10 claims over the 365 days prior to BUP initiation. Diagnoses were grouped into three categories: 1) risk associated with sex based on diagnosis of sexually transmitted infections (STIs): HBV, syphilis, chlamydia, or gonorrhea; 2) risk associated with injection drug use based on serious injection-related infections (SIRIs): hepatitis C, skin or soft tissue, infective endocarditis or septic arthritis) and 3) both STIs and SIRIs. See Appendix for more details. MarketScan data enabled observation of sex (male or female) and region (Northeast, North Central, South, West, and unknown). Age was operationalized as a linear measure in years. Race and ethnicity are not available through MarketScan data. Finally, to assess whether disenrollment might have contributed to low rates of observed HIV risk among BUP+PrEP episodes, we examined enrollment over the 365 days prior to BUP Day 1.

### Analysis

We used summary statistics to describe the distribution of covariates at the person-episode level. We described episodes according to those that did and did not involve PrEP during the treatment period. Due to the small number of episodes involving PrEP, we limited our analyses to descriptive statistics only.

## Results

We identified 124,536 BUP treatment episodes starting between 07/01/2015 and 10/30/2022 and excluded 796 episodes meeting criteria for HIV, leaving a total sample of 123,740 BUP treatment episodes representing 76,377 individuals. Of these episodes (Table 1), 104 involved PrEP (0.1%) at some point during BUP treatment (herein, BUP+PrEP episodes). Notably, 74 of these 104 BUP+PrEP episodes (71%) involved PrEP in the 30 days prior to BUP initiation. BUP+PrEP episodes were disproportionately male (89% compared with 63% for non-PrEP episodes) and from the Northeast (27% compared with 18% for non-PrEP episodes) or West (30% compared with 16% for non-PrEP episodes). Of the 104 BUP+PrEP episodes, 13% (13 episodes) exhibited a preceding HIV risk diagnosis.

**Table 1.**
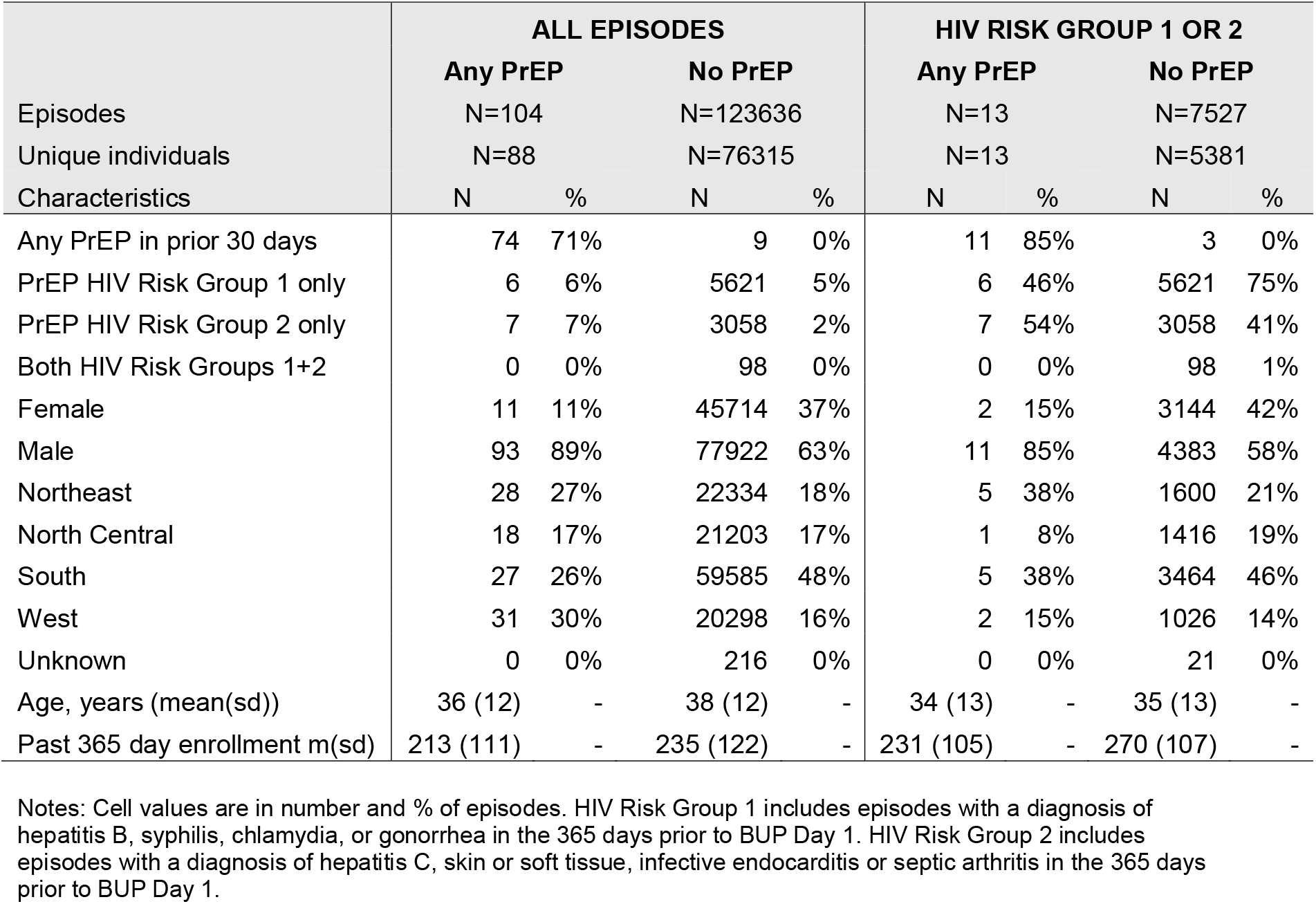
Characteristics of BUP treatment episodes with and without PrEP utilization.

Over 6% of the overall sample exhibited a preceding HIV risk diagnosis (7,540 episodes from 5,390 individuals); 98 episodes exhibited both an STI and SIRI diagnosis. Only 0.2% of BUP episodes with preceding HIV risk involved PrEP; none of the 98 episodes with both an STI and SIRI involved PrEP. Moreover, most HIV risk episodes that involved PrEP exhibited PrEP use prior to BUP initiation (11 of 13 episodes).

When considering episodes with HIV risk diagnoses, BUP+PrEP episodes were again disproportionately male (85% compared with 58% for non-PrEP episodes) and from the Northeast (38% compared with 21% for non-PrEP episodes). Episodes with an observed HIV risk diagnosis were enrolled more days on average than the full cohort, but past enrollment was higher among HIV risk episodes without PrEP (m=270 days; sd =107 days) than with PrEP (m=231; sd: 105).

## Discussion

In this large cohort of over 120,000 BUP treatment episodes, 0.1% involved PrEP during the first 63 days of BUP treatment. While not all patients with OUD seeking BUP treatment carry clinical indications for PrEP, among those with documented PrEP indications (6% of the sample), only 0.2% received PrEP during BUP treatment. These findings identify a major gap in PrEP for individuals with OUD and demonstrated HIV risk factors.

Our findings align with prior literature documenting significant PrEP underutilization for those at risk for HIV.^17–20^ This study advances the field by demonstrating an actionable opportunity to close this gap: many patients who seek BUP for OUD engage with care in the ambulatory setting where they can also receive screening for HIV risk and infection, and initiate PrEP as clinically indicated.^10^ This work is largely done by primary care clinicians^7,8^ who are well-trained and positioned at the front lines of prevention.^21,22^ Pharmacy claims cannot directly describe prescribing patterns by provider specialty. However, understanding these potential PrEP prescribing encounters could delineate a road map to close utilization gaps. To this end, empowering providers to incorporate PrEP into OUD treatment via system- and provider-level initiatives present an important opportunity to enhance HIV prevention among people with OUD.

Notably, most episodes involving PrEP received PrEP prior to BUP initiation. This finding suggests that PrEP prescribing could be a pathway to BUP treatment. For example, PrEP may reinforce patients’ health promotion goals and behaviors thereby enhancing motivation for OUD treatment. Future research and public health initiatives focused on OUD treatment and HIV prevention should consider these bidirectional paths.

Given that only 0.2% of BUP episodes with preceding HIV risk factors received PrEP, it is important to reflect on what strategies could help close this large gap. At the individual level, it is unclear what percentage of PrEP-eligible patients decline to initiate and what percentage are not offered to initiate PrEP. At the system-level, value-based care metrics and affordable care organization incentives could prompt health systems to develop workflows, best practices, or outreach teams to facilitate prescribing.^21–23^ Implementation research to bring evidence-based interventions such as BUP and PrEP into practice settings by targeting individual- and system-level determinants is needed.

This study was limited in several ways. First, while pharmacy claims confirm medication collection at the pharmacy, they cannot confirm precipitating events such as office visits. In the case of BUP treatment episodes, it is possible that some BUP prescribing occurred outside of office visits thereby overstating the number of missed opportunities to initiate PrEP. Second, we required brief continuous enrollment during the 30-day pre-period and 63 days of BUP treatment to expand sample inclusion given high rates of intermittent disenrollment in patients with OUD. As a result, we may have inadvertently included episodes with unobserved HIV diagnoses prior to BUP Day 1. Excluding episodes with HIV diagnoses or medications during the 63-day BUP treatment episode aimed to mitigate this limitation.

## Conclusion

In this study of over 120,000 BUP treatment episodes, 6% of episodes exhibited risk for HIV based on clinical diagnoses in the prior 365 days. However, only 0.2% of these episodes involved PrEP during the first 63 days of BUP treatment. These findings demonstrate substantial underutilization of PrEP among individuals with known risk factors for HIV despite participation in medication treatment for OUD. Optimizing HIV prevention within ambulatory BUP treatment represents an important opportunity to strengthen health care services for patients with OUD. In addition, given that most BUP episodes involving PrEP exhibited PrEP use prior to BUP initiation, initiatives aiming to improve OUD treatment and HIV prevention should consider these bidirectional paths.

## Data Availability

Data used and produced in the present study are not available for proprietary reasons.

## Notes

### Competing Interest Statement

The authors have declared no competing interest.

### Funding Statement

This study did not receive any specific grant from funding agencies in the public, commercial, or not-for-profit sectors. The following authors received salary support for their time: Tilhou (NIH K08DA058052); Assoumou (NIH/NIDA R01 DA058367); White (NIH NIGMS 5R35GM141821).

### Author Declarations

The study used HIPAA compliant de-identified data publicly available with purchase.

