## Appendix for "HIV pre-exposure prophylaxis among people initiating buprenorphine for opioid use disorder: a retrospective cohort study"

**Buprenorphine formulations**

1. 40225994 | buprenorphine 8 MG / naloxone 2 MG Sublingual Film [Suboxone] | 76160
2. 40225995 |buprenorphine 8 MG / naloxone 2 MG Sublingual Tablet [Suboxone]|18058
3. 40225990| buprenorphine 2 MG / naloxone 0.5 MG Sublingual Film [Suboxone]|4253
4. 42898504|buprenorphine 4 MG / naloxone 1 MG Sublingual Film [Suboxone]|3091
5. 19102739|buprenorphine 8 MG Sublingual Tablet [Subutex]|2952
6. 40225991|buprenorphine 2 MG / naloxone 0.5 MG Sublingual Tablet [Suboxone]|1520
7. 19102738|buprenorphine 2 MG Sublingual Tablet [Subutex]|599
8. 42898500|buprenorphine 12 MG / naloxone 3 MG Sublingual Film [Suboxone]|505
9. 793570|1.5 ML buprenorphine 200 MG/ML Prefilled Syringe [Sublocade]|420
10. 43532945|buprenorphine 5.7 MG / naloxone 1.4 MG Sublingual Tablet [Zubsolv]|355
11. 1133262|buprenorphine 8 MG / naloxone 2 MG Sublingual Tablet|233
12. 793482|0.5 ML buprenorphine 200 MG/ML Prefilled Syringe [Sublocade]|123
13. 45892568|buprenorphine 11.4 MG / naloxone 2.9 MG [Zubsolv]|39
14. 46287551|buprenorphine 2.9 MG / naloxone 0.71 MG Sublingual Tablet [Zubsolv]
15. 45892574|buprenorphine 8.6 MG / naloxone 2.1 MG Sublingual Tablet [Zubsolv]
16. 45774520  buprenorphine 4.2 mg / naloxone 0.7 mg buccal film [Bunavail]
17. 43532943|buprenorphine 1.4 MG / naloxone 0.36 MG Sublingual Tablet [Zubsolv]
18. 45776275|buprenorphine 2.1 MG / naloxone 0.3 MG Buccal Film [Bunavail]

**PrEP formulations**

emtricitabine/tenofovir alafenamide (NDC 61958200501)

emtricitabine/tenofovir disoproxil fumarate (NDC 00093760756)

cabotegravir (NDC 49702026423)

Oral cabotegravir was not included (NDC 49702024813)

**HIV risk indicators**:

Group 1: Injection drug use

Hep C: ICD-10: B18: ICD-9: 070.4, 070.5, 070.7
Soft tissue infection: ICD-10: L08.9; ICD9: 680-686
Infective endocarditis: ICD-10: I33; ICD9: 421, 424
infective arthritis: ICD-10: M00.8; ICD9: 711

Group 2: STIs

Chlamydia diagnosis: ICD-10: A74, A56  ICD-9: 099, 078, 079

Gonorrhea diagnosis: ICD-10: A54; ICD-9: 098

Hepatitis B infection: ICD-10: B19 ; ICD-9: 070.2 and 070.3, V02.6

Syphilis: ICD-10 A51, A52, A53; ICD-9: 091-097

**HIV Exclusions**

In line with existing literature on identifying PrEP in claims data,^1,2^ we chose to exclude episodes from individuals with possible HIV diagnoses based on presence of a HIV diagnosis (at least one outpatient encounter with an HIV diagnosis) or claim for an antiretroviral medication indicated for HIV based in the 365 days prior to Day 1 of buprenorphine treatment or during days 1-63 after buprenorphine initiation. To determine this medication list, we used all relevant components^3^ to identify monoproducts and combination products listed in Red Book for years 2014-2022. Component names are listed below. We included PrEP components in non-PrEP formulations: emtricitabine and tenofovir as monoproducts and oral cabotegravir.

HIV Diagnosis codes:

ICD-10: B20, B21, B22, B23, B24

ICD-9: ICD-9: 042 and V08.

HIV medications:

*Nucleoside Reverse Transcriptase Inhibitors*: abacavir, lamivudine, zidovudine, tenofovir, and emtricitabine

*Non-Nucleoside Reverse Transcriptase Inhibitors*: efavirenz, rilpivirine, nevirapine, etravirine, doravirine

*Integrase Strand Transfer Inhibitors*: elvitegravir, cobicistat, raltegravir, dolutegravir, cabotegravir

*Protease Inhibitors*: atazanavir, lopinavir, ritonavir, darunavir, fosamprenavir, tipranavir

*Fusion Inhibitors*: enfuvirtide

*CCR5 Antagonists*: [maraviroc](https://clinicalinfo.hiv.gov/en/drugs/maraviroc/patient)

*Attachment Inhibitors*: fostemsavir

*Post-attachment Inhibitors*: ibalizumab-uiyk

*Capsid Inhibitors*: lenacapavir

*Pharmacokinetic enhancers*: cobicistat
